# How public health decision-makers operationalise wastewater surveillance: a multi-region qualitative study

**DOI:** 10.64898/2026.05.14.26353119

**Authors:** Sana Zakaria, Henry H. Willis, Cindy R. Friedman, Mukhlid Yousif, Laura Faherty, Natalie Knox, Kerrigan McCarthy, Casey Aveggio, Derek Roberts, Adeline E. Williams, Saskia Popescu, Monica Nolan, Lionel Gresh, Jairo Andres Mendez Rico

**Affiliations:** RAND Europe, Cambridge, UK; RAND Corporation, Santa Monica, CA, USA; Friedman Global Health & Biosecurity, LLC, GA, USA; Centre for Vaccines & Immunology, National Institute for Communicable Diseases, National Health Laboratory Service, Johannesburg, South Africa; National Microbiology Laboratory, Canada; Promoting Health4All Pty Ltd; Infectious Hazard Management Unit, Health Emergencies Department, Pan American Health Organization, Washington DC, USA

## Abstract

**Background:** Wastewater and environmental surveillance (WES) expanded rapidly during the COVID-19 pandemic and is increasingly proposed for routine public health use across a broader range of pathogens. Yet empirical evidence on how decision-makers judge when WES is actionable, how it integrates with existing surveillance, and how its role varies across resource and epidemiological contexts remains limited.

**Methods:** We conducted three structured tabletop exercises (TTXs) at regional Global Wastewater Surveillance Consortium (GLOWACON) meetings in Singapore, Ethiopia, and Panama between March 2024 and May 2025, engaging more than 1,100 participants from over 60 countries spanning public health, government, research, industry, and international organisations. Standardised scenarios and decision prompts, covering respiratory, contact-transmitted, and vector-borne pathogens across multiple outbreak phases, elicited how participants prioritised, implemented, and responded to WES. Data from structured observation notes, participant worksheets, and post-exercise surveys were systematically analysed using a thematic qualitative approach to identify cross-cutting decision patterns and context-specific considerations across regions. This working paper has not been peer reviewed.

**Findings:** Four cross-cutting decision patterns emerged. First, WES was most actionable when it addressed defined surveillance gaps, particularly during early outbreak phases when clinical testing was limited or delayed. Second, decisions to initiate, scale, or de-escalate WES depended on disease severity, the availability of actionable interventions, and the completeness of existing surveillance, not on pathogen type. Third, participants consistently treated WES as complementary to, not a substitute for, clinical and epidemiological surveillance, with its role evolving over the course of an outbreak. Fourth, implementation considerations, including sewer infrastructure, resource constraints, tourism, and mass gatherings varied substantially by setting, while governance, data-sharing, and trust concerns recurred across all three regions.

**Interpretation:** The value of WES is determined less by pathogen-specific characteristics than by how it is embedded within decision-making frameworks in public health systems. These findings provide empirical evidence on how WES is operationalised across diverse global contexts and underscore an urgent need for clearer governance, integration, and prioritisation frameworks without which WES risks remaining an underutilised or inconsistently applied tool despite its demonstrated potential to strengthen pandemic preparedness and response.

**Funding:** This working paper was independently initiated and conducted within the Center on AI, Security, and Technology using income from operations and gifts and grants from philanthropic supporters. A complete list of donors and funders is available at www.rand.org/CAST. RAND clients, donors, and grantors have no influence over research findings or recommendations.

## Introduction

Wastewater and environmental surveillance (WES) has an established role in public health surveillance, most notably through its long-standing use in poliovirus monitoring^(1, 2)^ and its expanded application during the COVID-19 pandemic to complement clinical testing and case-based reporting.^(3)^ During COVID-19, WES provided community-level signals of transmission independent of healthcare access or testing availability, demonstrating its potential to address key surveillance gaps.^(4, 5)^

Interest in WES has since expanded to a broader range of pathogens of public health concern.^(6)^ At the same time, advances in laboratory and sequencing methods have increased the technical feasibility of WES across diverse settings.^(7, 8)^ However, despite this momentum, there remains limited empirical understanding of how WES is operationalised within public health systems in practice.^(7, 8)^ In particular, there is uncertainty around when WES is considered actionable, how it should be integrated with existing surveillance modalities, and how decision-makers weigh its role relative to other surveillance investments across epidemiological and resource contexts.

Most published WES research has focused on technical performance and proof-of-concept studies^(9)^, with far less attention to decision-making processes that govern deployment, scale-up, and sustained use. As WES is considered for more routine surveillance, understanding these processes is essential to ensure that WES contributes meaningfully to preparedness, response, and efficient use of limited public health resources.^(5)^

This study leverages the Global Wastewater Surveillance Consortium (GLOWACON), an international network of public health, academic, industry, and government stakeholders, to examine knowledge, attitudes, and practices related to WES implementation across diverse settings.^(10)^ Through structured tabletop exercises conducted at regional GLOWACON meetings in Singapore, Ethiopia, and Panama, we focus on decision frameworks to operationalise WES, including when and how to deploy it, how to translate WES findings into public health responses, and how to prioritise resources across surveillance platforms.

## Methods

### Study design and rationale

Structured TTXs were conducted to examine how public health decision-makers understand, prioritise, and operationalise WES within broader surveillance systems. Across three regional exercises, the study engaged more than 1,100 participants from over 60 countries, providing a substantive and rich cross-regional empirical datasets on WES decision-making. TTXs are qualitative, scenario-based exercises commonly used to elicit decision-making processes, identify thresholds for action, and explore trade-offs under uncertainty in public health preparedness.^(11, 12)^ The objective was not to simulate outbreak outcomes or evaluate technical performance, but to characterise decision logic related to when WES is considered actionable, how it is integrated with existing surveillance modalities, and how its role changes across outbreak phases and resource contexts.

### Scenario selection and disease rationale

Three tabletop exercises were conducted at regional meetings of GLOWACON, each centred on a distinct disease scenario. Scenarios were chosen to enable comparison of WES-related decision-making across respiratory, vector-borne, and contact-transmitted pathogens, and across regions with differing public health priorities and surveillance capacity.

The Singapore exercise focused on a highly pathogenic respiratory virus with zoonotic origin and potential human-to-human transmission, reflecting challenges related to early detection, importation risk, and escalation thresholds in a high-capacity setting. The Ethiopia exercise centred on a hypothetical novel poxvirus requiring notification under the International Health Regulations, designed to represent a contact-transmitted disease in an African context where surveillance systems face capacity constraints and international coordination plays a critical role. The Panama exercise focused on an arboviral disease with potential for endemic transmission, selected to examine decision-making for vector-borne surveillance in a Latin American setting where economic considerations, including tourism and mass gatherings, influence public health response.

Detailed scenario narratives and inject timelines are provided by exercise in Supplementary materials 1-3.

### Exercise structure and facilitation

Each TTX was conducted over approximately four hours and ran in parallel in-person and online streams to maximise participation across the global GLOWACON community. In-person participants worked in small, facilitated groups, followed by plenary discussions to synthesise perspectives across groups. Online participants contributed concurrently through structured digital instruments—a shared Mural collaborative whiteboard and online forms—that mirrored the prompts and worksheets used in the in-person sessions, enabling remote attendees to engage with the same scenario injects and decision domains in real time. Exercises progressed through a series of scenario injects that revealed new information over time, simulating the evolving nature of outbreak detection, response, and stabilisation. Facilitators guided discussion using standardised prompts but did not direct participants toward specific conclusions or optimal solutions. While the overall structure and core decision domains were consistent across exercises, contextual adaptations were made to ensure relevance to regional public health systems.

### Decision frameworks and discussion prompts

Discussion prompts were developed to elicit how participants reasoned about surveillance choices under uncertainty and resource constraints. Across all exercises, prompts focused on a common set of decision domains, including:

1. triggers for initiating WES;
2. conditions for scaling WES up or down;
3. integration of WES with existing clinical, laboratory, epidemiological, and, where relevant, airport surveillance systems;
4. prioritisation of limited surveillance resources; and
5. governance, data sharing, and communication considerations.

Standardised decision prompts are provided by exercise in Supplementary materials 1-3.

### Data collection

Data sources spanned both participation streams and included: structured observation notes taken by facilitators during in-person small-group and plenary discussions; completed participant worksheets from in-person groups; contributions to the shared Mural collaborative whiteboard used by online participants; and individual responses to the online forms completed by remote attendees. No personal identifying information was collected. In-person data were captured at the level of small-group discussion, while Mural and online-form data were captured at the level of individual or self-organised online-group inputs; all sources were consolidated for analysis across the three exercises.

### Data analysis

Data were analysed using a thematic qualitative approach with inductive coding. Two members of the research team independently coded all facilitator observation notes, participant worksheets, Mural board contributions, and online-form responses, allowing themes to emerge from the data rather than from a predefined codebook. Discrepancies between coders were resolved through discussion to converge on a shared coding, which was then refined through iterative cross-comparison across the three regional exercises to distinguish cross-cutting decision patterns from context-specific considerations. Analysis focused on identifying the conditions under which WES was considered actionable, the factors shaping decisions to initiate or scale WES, and how its perceived role shifted across outbreak phases and settings. The analysis was descriptive and comparative in nature and did not seek to test predefined hypotheses. Initial analyses of themes are provided by exercise in Supplementary materials 1-3.

## Results

### Participants

Between March 2024 and May 2025, three structured TTXs were conducted at regional GLOWACON conferences in Singapore, Ethiopia, and Panama, representing diverse regions, and engaging over 1,100 participants from more than 60 countries from the global wastewater surveillance community (Table 1). The participants ranged from across public health, research, government, industry, and international organisations. Each TTX possessed unique characteristics, including geographic setting, disease scenario, primary surveillance challenges, and key decision domains explored (Table 2). We first describe findings that were consistent across all three exercises and then highlight themes that were specific to particular regional contexts. Table 3 summarises which themes were most prominent in each exercise, distinguishing cross-cutting findings from context-specific observations.

**Table 1.** Countries and sectors most represented at the three GLOWACON meetings.

| Conference site | Singapore | Ethiopia | Panama |
| --- | --- | --- | --- |
| Number of registrants | 243 onsite | 110 onsite | 94 onsite |
|  | 156 remote | 204 remote | 306 remote |
| Countries with majority representation<sup>1</sup> (number of participants) | Singapore (92) | US (31) | US (55) |
|  | US (26) | Ethiopia (22) | Brazil (22) |
|  | India (23) | Germany (22) | El Salvador (19) |
|  | UK (19) | UK (15) | UK (19) |
|  | Indonesia (17) | South Africa (14) | Argentina (17) |
| Sectors most represented (number of participants) | Research institutes (61) | Research institutes (102) | Research institutes (122) |
|  | Industry (26) | Public Health Organizations (74) | Public Health Organizations (83) |
|  | Public Health Organizations (25) | National Government (32) | National Government (60) |
|  | National Government (18) | International Organizations (31) | International Organizations (36) |

**Table 2.** Overview of GLOWACON Tabletop Exercises. WES = wastewater and environmental surveillance; IHR = International Health Regulations

| Characteristics | Asia TTX (Singapore) | Africa TTX (Ethiopia) | Latin America TTX (Panama) |
| --- | --- | --- | --- |
| Number of countries | 59 | 60 | 67 |
| Primary focus | Decision frameworks for initiating WES | Application of WES insights to guide public health response | Prioritisation frameworks for selecting surveillance modalities |
| Disease scenario | Highly pathogenic respiratory virus with zoonotic origin and potential human-to-human transmission | Novel poxvirus with close-contact transmission requiring IHR notification | Vector-borne virus with potential for sustained endemic transmission |
| Scenario progression | From animal detection to confirmed human-to-human transmission | From initial WES detection to widespread community outbreak | From initial cluster to amplification during a regional mass gathering event |
| Key themes that emerged during decision-making and escalation | - Thresholds and triggers for initiating WES<br>- Criteria for escalation and public health action | - Activation of response systems<br>- Coordination across international and regional partners<br>- Data sharing | - Selection between wastewater and vector surveillance modalities<br>- Managing surveillance during tourism and travel events |

<sup>1</sup> Participants from other countries were in attendance beyond the ones listed in Table 1.

|  |  |  |  |
| --- | --- | --- | --- |
| **points** | - Resource mobilisation and cross-sector communication | protocols and capacity strengthening | - Coordination across urban and rural jurisdictions |
| **Resource context** | High-income setting with existing infrastructure | Low-income setting with capacity constraints | Mixed-resource setting with tourism dependency |

**Table 3.**
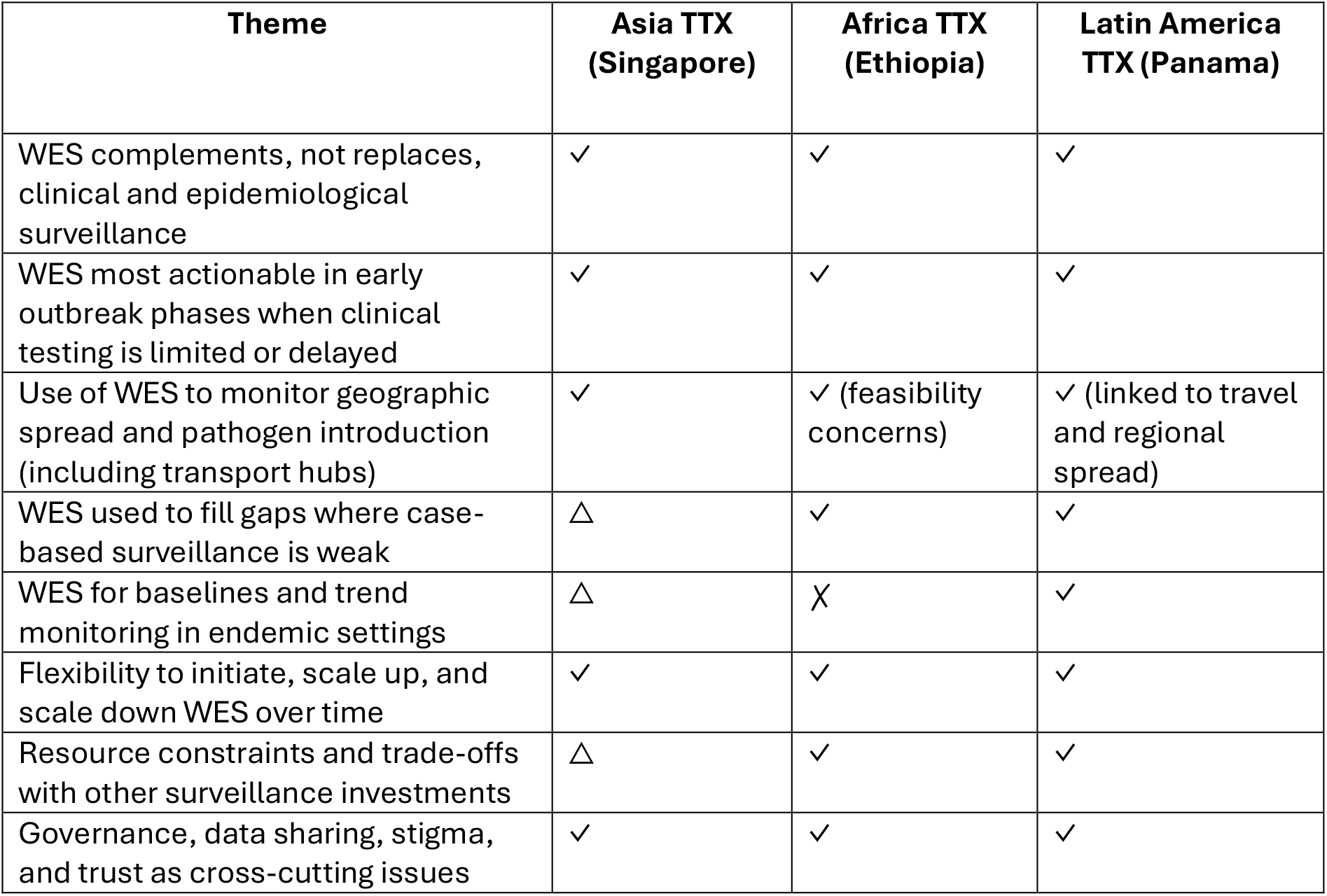
Summary of key WES decision-making themes by tabletop exercise.

| Theme | Asia TTX<br>(Singapore) | Africa TTX<br>(Ethiopia) | Latin America<br>TTX (Panama) |
| --- | --- | --- | --- |
| WES complements, not replaces, clinical and epidemiological surveillance | ✓ | ✓ | ✓ |
| WES most actionable in early outbreak phases when clinical testing is limited or delayed | ✓ | ✓ | ✓ |
| Use of WES to monitor geographic spread and pathogen introduction (including transport hubs) | ✓ | ✓ (feasibility concerns) | ✓ (linked to travel and regional spread) |
| WES used to fill gaps where case-based surveillance is weak | △ | ✓ | ✓ |
| WES for baselines and trend monitoring in endemic settings | △ | ✗ | ✓ |
| Flexibility to initiate, scale up, and scale down WES over time | ✓ | ✓ | ✓ |
| Resource constraints and trade-offs with other surveillance investments | △ | ✓ | ✓ |
| Governance, data sharing, stigma, and trust as cross-cutting issues | ✓ | ✓ | ✓ |

### Conditions under which WES was considered actionable

Across all three exercises, participants consistently described WES as actionable when it addressed clearly defined surveillance gaps and when outputs could plausibly inform subsequent public health actions (Table 3). Across exercises, actionability was not treated as an intrinsic property of WES data, but as contingent on epidemiological context, disease characteristics, and the availability of complementary information.

### Early population-level detection

Across all exercises, participants reported that WES was most actionable during early outbreak phases, particularly when clinical testing was limited, delayed, or unevenly distributed (Table 3). In the Africa region exercise, groups emphasised WES as a means of detecting transmission that might otherwise go unobserved because of barriers to healthcare access, stigma associated with testing, or limited laboratory capacity. In these contexts, WES was viewed as a trigger for heightened vigilance and targeted follow-up rather than as a stand-alone signal for intervention.

### Monitoring geographic spread and pathogen introduction

Across all exercises, participants discussed the use of WES to monitor pathogen introduction and geographic spread, especially through transport hubs, although the perceived feasibility and priority of this application varied by setting (Table 3). In the Asia region exercise, many groups supported wastewater monitoring at airports as a means of identifying importation risk and contextualising early signals of transmission. In contrast, participants in Africa TTX expressed more caution, balancing the potential surveillance benefit against concerns about feasibility and economic or reputational implications. In the Latin America setting, there was stronger consensus that transport hub monitoring could support situational awareness for returning travellers and regional spread, particularly during periods of increased population movement.

### Variant and strain detection

Participants across exercises identified genomic analysis of wastewater as a potentially important function but differed in how it influenced decision-making. In Singapore, groups prioritised variant detection once sustained transmission was established, describing WES as a tool to complement clinical sequencing and track changes over time. In lower-capacity contexts, participants emphasised the need for confirmatory clinical data before acting on variant signals, noting that interpretation without supporting epidemiological context could be challenging.

### Establishing baselines for endemic diseases

In the Panama exercise, participants highlighted the role of WES in establishing baseline levels for endemic pathogens, allowing deviations from expected patterns to be detected. Rather than serving as an early warning of emergence, WES was viewed as a tool for monitoring trends over time and supporting decisions about when to intensify or relax other surveillance activities. This use case was discussed less frequently in the Singapore exercise and was largely absent from Ethiopia, reflecting differences in disease priorities and surveillance objectives, and indicating that baseline monitoring was a context-specific theme primarily associated with the Panama exercise.

### Flexibility and responsiveness over time

Participants frequently emphasised the ability to adjust WES intensity over time as a key consideration. In the Singapore exercise, groups described an explicit transition point: initial hesitancy to implement broad WES in the absence of confirmed human transmission shifted to unanimous support once human-to-human transmission was established in the scenario. As the scenario progressed further, participants recommended reducing sampling frequency and narrowing geographic coverage, reflecting changing cost–benefit considerations as clinical surveillance became more informative.

### Disease severity

Disease severity strongly influenced willingness to invest in intensive WES. Participants described higher tolerance for resource-intensive surveillance when diseases were perceived as severe or associated with high uncertainty. In contrast, for lower-severity or endemic conditions, WES was more often framed as a supplementary or sentinel tool rather than a primary surveillance modality.

### Availability of actionable interventions

Participants repeatedly linked the usefulness of WES to the availability of interventions that could plausibly follow detection. Where vaccines, therapeutics, or targeted public health measures were available, WES signals were viewed as more actionable. In scenarios where few response options existed, participants expressed greater caution, noting the risk of generating signals that could not be translated into meaningful action.

### Completeness and timeliness of existing surveillance systems

The perceived added contribution of WES depended on the strength of existing surveillance. In high-capacity settings, participants described WES as confirmatory or complementary, supporting trend analysis and validation of clinical data. In lower-capacity settings, WES was more often framed as a gap-filling tool, particularly where case-based surveillance was delayed or incomplete, whereas in higher-capacity settings it was described more as a confirmatory or complementary source of information (Table 3).

### Integration with other surveillance systems

Participants consistently described WES as one component within broader surveillance systems rather than as a stand-alone approach. Across exercises, groups discussed using WES signals to guide downstream surveillance activities, including targeted clinical testing, expanded laboratory investigation, or enhanced vector surveillance. In Panama, participants described how wastewater signals could prompt intensified vector surveillance and targeted polymerase chain reaction (PCR) testing in specific areas, while in Ethiopia, groups discussed using WES findings to prioritise limited clinical testing resources. In Singapore, participants emphasised bidirectional integration, with clinical data guiding WES sampling strategies and wastewater trends providing additional context for interpreting case data. These discussions highlighted the importance of coordination across surveillance streams to avoid duplication and to support proportionate responses.

### Infrastructure and data integration

Across exercises, participants emphasised the importance of established sampling, laboratory, and data management infrastructure. In Ethiopia and Panama, groups noted challenges related to limited sewer coverage and highlighted the need for alternative sampling strategies. Participants in all settings emphasised the value of integrated data platforms that combined WES with clinical and epidemiological data to support timely interpretation.

### Resource and financing considerations

Participants frequently discussed resource constraints and trade-offs between WES and other public health investments, with these concerns most prominent in the Africa and Latin America exercises. Several groups noted that investments initially made for single-pathogen applications, such as SARS-CoV-2 or poliovirus surveillance, could be leveraged for monitoring additional pathogens, influencing how resource allocation decisions were framed. Nonetheless, participants emphasised the need for clarity about when WES offered sufficient added benefit to justify continued investment.

### Governance, ethics, and trust

Concerns related to data sharing, communication, and potential stigma were raised across exercises. Participants emphasised that trust between institutions, across sectors, and with communities was critical for sustained WES implementation. In particular, groups in Ethiopia and Panama highlighted the importance of avoiding punitive responses to outbreak detection that could discourage transparency and data sharing.

## Discussion

Drawing on structured exercises with more than 1,100 participants from over 60 countries, this study provides empirical evidence on how WES is currently understood and operationalised at the interface between technical capability and public health decision-making. Rather than identifying novel technical functions of WES, the findings demonstrate that its contribution to public health surveillance is primarily shaped by how decision-makers frame actionability, proportionality, and integration within existing systems. Across diverse disease contexts and resource settings, WES was not treated as a universally applicable solution, but as a contingent tool whose relevance depends on epidemiological uncertainty, response objectives, and competing surveillance priorities.

A central implication of these findings is that the effectiveness of WES is determined less by pathogen-specific characteristics than by the presence of decision frameworks that guide when and how surveillance data are used. Existing applications of WES, most notably for poliovirus and COVID-19 surveillance, have demonstrated its technical feasibility and utility under specific conditions.^(13)^ Beyond these settings, formal guidance on how WES should be deployed across different pathogens, outbreak phases and surveillance environments remains limited.^(14)^ The decision patterns observed here highlight a persistent gap between the generation of wastewater data and the translation of those data into proportionate public health responses, suggesting that future investments should prioritise decision frameworks and governance mechanisms alongside technical development.

The findings also point to an important shift in how WES is conceptualised within public health systems. Rather than being viewed solely as a pathogen-specific surveillance tool, WES increasingly appears to be understood as a platform capability that can support multiple surveillance objectives over time.^(15)^ This framing has implications for resource allocation, particularly in contexts where investments made for one disease (such as SARS-CoV-2 or poliovirus) can be leveraged for broader preparedness and response. At the same time, the platform perspective does not remove the need for clear prioritisation criteria, especially in resource-limited settings where surveillance investments must be balanced against other essential public health functions.

Another key implication relates to equity and sustainability. While interest in WES is global, the capacity to implement and sustain surveillance varies substantially across settings.^(15, 16)^ The findings reinforce broader systemic concerns that, without deliberate attention to financing, infrastructure, and governance, new surveillance systems could exacerbate existing inequities in surveillance capacity. Short-term pilot projects and emergency funding, while valuable for demonstrating feasibility, are unlikely to support long-term integration into public health systems. Aligning WES implementation with broader health system strengthening efforts and ensuring that data-sharing arrangements do not penalise countries for transparent reporting, will be critical for equitable and sustainable use. Notably, participants in lower-resource settings described WES as a relatively low-cost sentinel layer that could help direct where more resource-intensive clinical and laboratory surveillance is deployed, a framing with potential to ease, rather than reinforce, surveillance inequities, provided governance and financing structures support sustained use.

These exercises provide empirical evidence on how practitioners and policymakers currently interpret and apply WES within public health systems. By engaging those already working in WES across diverse regions, the exercises capture emerging expectations about how these approaches may be integrated into routine practice.

While this perspective may reflect some optimism about the role of WES, it also highlights how the field is evolving from emergency-driven applications toward more sustained use. In this sense, the findings complement formal evaluations and policy frameworks by illustrating how surveillance tools are interpreted and prioritised in real world settings, particularly where guidance remains limited.

Altogether, these findings suggest that the future contribution of WES to global public health will depend not only on continued methodological advances, but on the development of clear, context-sensitive frameworks for decision-making, integration, and governance. Without such frameworks, WES risks remaining an underutilised or inconsistently applied tool despite its demonstrated potential.

### Policy implications

#### Clarifying the role of WES within integrated surveillance systems

Policies should explicitly position WES as one component within integrated surveillance frameworks, rather than as a stand-alone solution. Guidance is needed on the conditions under which WES is most likely to inform public health action, including disease severity, outbreak phase, availability of interventions, and the completeness of existing surveillance systems. Clear criteria for initiating, scaling, and de-escalating WES would support proportionate use and more transparent resource allocation.

#### Supporting cross-sectoral coordination for surveillance decision-making

Participants highlighted the need for clearer coordination across public health, laboratory, environmental, and, in some contexts, biosecurity stakeholders. Policy guidance that defines roles, responsibilities, and data-sharing arrangements across sectors could support more effective interpretation and use of WES findings, particularly where surveillance signals inform broader response decisions such as resource prioritisation, risk communication, or deployment of countermeasures.

#### Framing WES investment as a platform capability

Resource allocation decisions would benefit from policy guidance that recognises WES infrastructure as a platform that can support multiple pathogens over time, rather than as a series of pathogen-specific investments. Such framing may improve sustainability and preparedness, particularly where infrastructure developed for diseases such as SARS-CoV-2 or poliovirus can be leveraged for additional surveillance objectives. However, this approach should be accompanied by explicit prioritisation criteria to avoid displacing other essential public health functions, especially in resource-limited settings.

#### Embedding governance and equity considerations into WES policy

Clear governance frameworks are needed to guide data ownership, sharing, and communication of WES findings. Policies should address risks of stigma or economic harm associated with outbreak detection and reporting, and promote trust by ensuring that countries and communities are not penalised for transparent surveillance. International and regional partners can support these efforts, but long-term effectiveness will depend on alignment with national priorities and sustainable financing mechanisms.

### Limitations

Several limitations should be considered. First, TTXs elicit stated decision logic in hypothetical scenarios rather than observed deployment behaviour; while this design is well suited to surfacing reasoning under uncertainty, it cannot establish what decision-makers would do under real-world political, financial, and operational pressures. Second, data sources differed in granularity across the in-person and online streams: in-person inputs were captured primarily at the level of small-group discussion, while online inputs included individual-level form responses and Mural contributions. This mixed granularity supported broad consolidation across participation modes but limits formal inference about within-group divergence and the influence of participant background; some participants also attended multiple TTXs, and these effects were not modelled. Third, participant selection was not random but comprised individuals already engaged with the GLOWACON network, potentially introducing selection bias toward those more favourable to WES approaches. Fourth, while participants represented over 60 countries, regional voices were not evenly distributed across exercises, and findings may not generalise to settings outside the GLOWACON community. Finally, the exercises primarily focused on infectious disease surveillance applications, with limited exploration of other potential uses, including antimicrobial resistance monitoring and biological and chemical threat detection. These limitations can be addressed through prospective programme evaluations that compare stated decision logic with actual deployment decisions, by engaging broader groups of policymakers and public health officials, and by conducting exercises using a broader range of scenarios and threat types.

## Supporting information

Panama Event S1

Ethiopia Event S2

Singapore Event S3

## Author contributions

Sana Zakaria and Henry Willis conceptualised and outlined the working paper. Derek Roberts and Casey Aveggio performed thematic analysis. Sana Zakaria, Adeline E. Williams, Henry Willis, and Derek Roberts reviewed and edited the working paper. Sana Zakaria, Henry H. Willis, Cindy R. Friedman, Mukhlid Yousif, Laura Faherty, Natalie Knox, Kerrigan McCarthy, Monica Nolan, Lionel Gresh, Jairo Andres Mendez Rico, Adeline E. Williams, and Saskia Popescu contributed to exercise design and execution.

## Competing Interests

The authors declare no competing interests

## Ethics statement

This study involved structured group-discussion exercises with public health, government, research, industry, and international-organisation professionals attending regional GLOWACON meetings, delivered through parallel in-person and online streams. Informed consent for participation in the regional GLOWACON exercises was obtained separately by the GLOWACON organising committee at the point of conference registration; all participants engaged voluntarily. The study is exempt from full human subjects research review on the grounds that data were captured at the level of small-group discussion (in-person) or as anonymised individual contributions via the Mural collaborative whiteboard and online forms (online), with no personal identifying or sensitive personal information collected and with the dataset not capable of identifying individual participants.

## Data availability

De-identified, group-level summary data supporting the findings of this study including thematic syntheses of facilitator observation notes, participant worksheets, and post-exercise survey responses are available from the corresponding author on reasonable request. The full scenario narratives, decision prompts, and inject timelines used in each of the three exercises are provided in Supplementary Materials 1–3.

## Acknowledgments

The authors would like to acknowledge Jean Pierre Musabyimana RBC, Bernd Gawlik, Angela Tessarolo, Anastasia Koutsolioutsou, and Rosa Maria Torres for support to this manuscript.

## Center on AI, Security, and Technology

RAND Global and Emerging Risks is a division of RAND that delivers rigorous and objective public policy research on the most consequential challenges to civilization and global security. This work was undertaken by the division’s Center on AI, Security, and Technology, which aims to examine the opportunities and risks of rapid technological change, focusing on artificial intelligence, security, and biotechnology. For more information, contact.

*RAND working papers are intended to share early insights and solicit informal peer review. This working paper has been approved for circulation by RAND but has not been peer reviewed or professionally edited or proofread. Review comments are welcomed at the Rxiv version of this paper. This working paper can be quoted and cited without permission of the author, provided the source is clearly referred to as a working paper. This working paper does not necessarily reflect the opinions of RAND’s research clients and sponsors*.

## Abbreviations

*WES*: *Wastewater and environmental surveillance*
*TTXs*: *Tabletop exercises*
*GLOWACON*: *Global Wastewater Surveillance Consortium*
*PCR*: *Polymerase chain reaction*

## Notes

### Competing Interest Statement

The authors have declared no competing interest.

### Author Declarations

Informed consent for participation in the regional GLOWACON exercises was obtained separately by the GLOWACON organising committee at the point of conference registration; all participants engaged voluntarily. The study is exempt from full human subjects research review on the grounds that data were captured at the level of small-group discussion (in-person) or as anonymised individual contributions via the Mural collaborative whiteboard and online forms (online), with no personal identifying or sensitive personal information collected and with the dataset not capable of identifying individual participants.

