## Supplementary material for "How public health decision-makers operationalise wastewater surveillance: a multi-region qualitative study": Panama Event S1

*Supplementary Data 1: Panama GLOWACON Tabletop Event*

### Event Scenario & Prompts

Today's exercise takes place in fictitious non-contiguous countries in the Americas. "Riofiebre virus" is fictitious, although it has characteristics of existing pathogens. This scenario takes place in 2025-2026.

**Latin America context:**

Country 1 is **San Pedro**, a South American country that is highly sewered. About 75% of people are connected to a sewage network system while those in more rural areas either rely on septic tanks or lack sanitation infrastructure. WES is being done at a national level by research institutions and the Ministry of Health for polio and SARS-CoV-2.

Country 2 is **Val Verde**, a Central American country with mixed sewage systems. Some large cities have partially convergent sewage systems, while only half of those in rural areas have access to either a piped sewer system or a septic tank system. Limited WES is being conducted for polio with support from NGOs and a handful of international research institutions.

Country 3 is **Tropico**, a Caribbean island nation with a vibrant tourism industry, international airport, and cruise ship port. There are minimal sewer systems (less than 20% of people are covered), and most places rely on septic tanks. WES is not typically performed, although some research institutions have experimented with it for SARS-CoV-2, and a system had been put in place during the COVID-19 pandemic to have samples shipped and sequenced at a PAHO/WHO reference laboratory. Several international research institutions work in Tropico because of its vibrant biodiversity and perform One Health surveillance.

**North America context:**

Country 4 is **Pacifica**, which has highly networked sewer systems and ongoing government-run WES in communities and airports.

**Base Scenario:**

It's September 2025, and a research institution performing vector surveillance in remote areas of the South American country of San Pedro detects riofiebre virus in a sizable number of mosquitoes. Riofiebre is a mosquito-borne illness known to be endemic in the tropical rainforest of San Pedro, and the zoonotic reservoirs are believed to be sloths, non-human primates, and birds. It is transmitted by multiple vectors, including mosquitoes, with the primary vector being *Culicoides paraensis* midges that have a wide geographical distribution across the Americas. The species of mosquito in which the virus was detected was known to maintain the sylvatic virus cycle (circulation of virus between mosquitoes and animals, without involving humans).

### *Supplementary Data 1: Panama GLOWACON Tabletop Event*

Although riofiebre outbreaks in humans had been reported in San Pedro and other countries throughout central and South America in the past (meaning, there had been sustained human-vector-human transmission), they are uncommon, so the virus is not typically screened for in clinical settings. Approximately 40% of infected individuals also remain asymptomatic and never get diagnosed with the virus. There are no available treatments.

Two months later, in November 2025, a research team from an externally funded NGO is doing metagenomic analysis of wastewater samples throughout San Pedro, and they identify riofiebre virus in wastewater samples collected at sites located in three mid-sized cities around the tropical rainforest. The WES detection occurs before any clinical cases have been identified.

Fast forward to the next year, in March 2026, and 20 patients visit a major clinic in the capital city of San Pedro with fever and other mild non-specific symptoms (headache, malaise, and nausea). They had all been part of a group of 100 people who recently traveled together on a two-week expedition to a tropical rainforest in the country. The patients began feeling unwell once they returned home. Because of their travel history, clinicians order RT-PCR and antibody tests for major arthropod-borne (arbo) viruses endemic to the area – Zika, dengue, and chikungunya viruses. The RT-PCR tests come back negative, although several of the patients had dengue- and Zika-positive IgG antibodies. Clinicians recommend supportive care to control fever and advise the patients to return to the clinic if symptoms worsen. All patients recover in one to two weeks.

Now it's May 2026, and several patients who were among the group of travelers to the rainforest begin experiencing a recurrence of symptoms (high fevers, severe headache, myalgia, and nausea) and come back to the clinic. One of the clinicians attending to the patients had recently read about riofiebre in a research publication on a historical case study, prompting them to order a pan flaviral RT-PCR test, which includes testing for riofiebre. Blood and urine for all returning patients are positive for riofiebre.

These results prompt the clinician to reach out to the local public health authorities and report that several recent travelers from the San Pedro tropical forest are being diagnosed with riofiebre virus. Epidemiologists then contacted, interviewed, and tested PCR and antibody titers from the additional 80-90 travelers from the trip that had either remained asymptomatic or who had initially presented with fever to the clinic but had not experienced a relapse of symptoms. From this investigation, the epidemiologists find that ~40 additional individuals are either RT-PCR positive for riofiebre or have riofiebre-specific IgG antibodies.

**Base Scenario Prompts:**

1. What was the significance of the WES finding and the level of concern it may warrant from a PH perspective?
    - What could the sources of the WES signal be? What tools could you develop to detect if the source is human or non-human?
    - What characteristics of riofiebre virus contribute to the significance of the WES finding?

- How might wastewater detection inform other surveillance, public health, and clinician action at this point?
- What would you have done with the WES insights on riofiebre detection before any clinical cases were identified?
- What information should be shared about the WES finding, vector surveillance finding, and cluster of cases?
- Furthermore, what information-sharing systems/relationships in public health agencies are responsible for sharing this information and to whom?

2. What should San Pedro public health officials consider doing next, now that 40 additional individuals from the tour group show laboratory evidence of riofiebre infection?
    - What is the "menu" of possible public health responses that could be considered?
    - Which ones are most important to pursue now, with existing information? Which ones need to be deferred until more information is available? What additional information is needed to move forward with other public health responses?
    - Would you recommend additional confirmatory testing by the existing WES sites if they are run by research institutions, NGOs, or private entities?
    - Are there partnerships that may be required for expanded testing?
    - Should surveillance be expanded to additional sites or other biosurveillance strategies?
    - What collaborative or integrative surveillance measures should be implemented at this point? (please explore sample type, geographic region, etc.)
3. Explain the roles of the NGO doing WES in San Pedro, the San Pedro public health institute. and PAHO/WHO at this point in the investigation?
    - What are key considerations for a successful joint investigation?

**Inject 1:**

Now it's July 2026. Detection has grown beyond WES, and clinical cases of riofiebre are rising in San Pedro. The San Pedro Ministry of Health is now reporting six possible cases of maternal-to-child transmission of riofiebre disease during pregnancy. A 2024 paper studying outcomes from previous riofiebre outbreaks noted possible associations to congenital malformations among children of mothers infected during their second trimester of pregnancy. It is also now known that neuroinvasive disease occurs in as many as 5% of patients.

At this time, many countries throughout the Americas, including San Pedro, are preparing to host a month-long 2026 International Football Championship in August, which leads health officials in San Pedro to increase syndromic surveillance and issue health alerts for riofiebre virus to clinicians and public health professionals across the region in anticipation of increased international travel. The health alerts are also distributed to two countries gearing up to host the International Football Championship: Val Verde in Central America and Pacifica in North America. Both countries are considering whether to expand public health preparedness (e.g. clinical alerts) and surveillance efforts targeted for the riofiebre virus. Unlike Pacifica, Val Verde is also experiencing a severe dengue outbreak, stressing the public health and healthcare infrastructure. The National Public Health Laboratory Network of Val Verde, which was already on alert due to the upcoming tournament, is now activated to identify, plan, and report a strategy for riofiebre surveillance once the games begin.

**Inject 1 Prompts:**

1. What value, if any, would WES have in San Pedro, Val Verde, and Pacifica at this point?
    - List specific factors that might affect the decision to conduct WES
    - Should any of these countries consider integrating other surveillance modalities (including vector surveillance) into the surveillance network, if it might come at the cost of expanding WES?
    - How do multiple circulating arboviruses impact your decision about surveillance?
2. What potential problems (and solutions) for implementing, sustaining, and/or expanding WES in San Pedro and Val Verde should be considered?
    - What capacity bottlenecks?
    - What infrastructure issues?
    - What can be done to resolve issues?
    - Should Pacifica initiate WES targeted to riofiebre? Why or why not?
3. How could you best integrate WES sampling collection, analysis, and resulting data with existing surveillance systems and laboratory networks? What would collaboration with national and regional disease control centers look like?
4. What would the communication plan be for travelers, pregnant women, and clinicians, given what you know so far?
    - How would you communicate WES signals and the uncertainty of riofiebre circulation based on WES in your risk communication messaging to various groups (e.g. PH, policy, clinicians, general public, pregnant women)
    - Would you feel comfortable basing your risk assessment on the WES data available so far, or what additional data would you need for PH risk messaging?
5. Would there be a value for doing aviation wastewater surveillance at this point- if so, which countries would benefit from this?
6. What should the regional lab network consist of and what should their priorities be for establishing a response plan, with consideration to the stressed systems and riofiebre cases load.

**Inject 2:**

Now it's mid-August 2026, and the International Football Championship is well underway. Vector season is peaking in Val Verde and Pacifica but is winding down in most of San Pedro. Val Verde is experiencing major outbreaks of both riofiebre and dengue. Pacifica has detected riofiebre in both WES and in vector populations, raising concerns about local transmission, but only  a handful of clinically confirmed riofiebre cases have been detected in Pacifica, all of which were travel associated.

*Supplementary Data 1: Panama GLOWACON Tabletop Event*

Fortunately, two world-renowned institutes in North and South America have developed a new 2-dose riofiebre vaccine that has been approved for use in Val Verde. The Val Verde Ministry of Health begins rapidly deploying the vaccine, but low turnout, especially for the second dose of the vaccine, may limit its impact; many pregnant individuals are also voicing hesitancy to take it since it is a new vaccine.

Meanwhile, at a few WES sites, a research institute in Val Verde began using a novel wastewater sample processing method that enables higher efficiency and higher depth sequencing of both dengue & riofiebre. Based on this success, a global development organization sets aside a large amount of money for metagenomic sequencing of WES across Latin America, including in San Pedro & Val Verde. Results of this effort suggest that multiple genotypes of Riofiebre are circulating in different regions.

Meanwhile, as San Pedro and Val Verde continue to welcome travelers for the last week of the 2026 International Football Championship, to riofiebre and its clinical sequelae have made international news.

Elsewhere, the vacation hub of Tropico is in its peak tourist season. The island is expected to have record numbers of visitors due to the International Football Championship, which is set to attract over 2.5 million football fans over the one-month course of the games, including 1 million international visitors. Tourism is the major source of income for Tropico, which was severely impacted during the COVID-19 pandemic.

**Inject 2 Prompts:**

1. What types of public health response actions could be triggered by WES findings both nationally and globally?
    - Consider investigation and enhanced surveillance
    - Consider engagements across government and research stakeholders
    - Consider public communication
    - Consider international stakeholder engagement
    - Consider epidemiology (including molecular epidemiology) of riofiebre and dengue
    - Consider cross border issues that may be triggered by the WES findings
2. What additional WES research should be encouraged to optimize WES methods for riofiebre & dengue detection and to better understand its molecular biology and epidemiology?
    - Consider the virus's persistence and viability in wastewater
    - Consider impact of medical and non-medical interventions, including vector control and vaccine roll out & evaluation

4. Should smaller, regional countries without cases and minimal WES systems, establish testing for riofiebre? Why or why not? What would prompt stronger recommendations for them to implement WES?
5. Should there be additional actions surrounding the football championship? The health ministries of San Pedro and Val Verde have been coordinating with the championship

safety officials, but are now asking if they should expand notifications, prevention protocols, and surveillance networks, including WES, traveler (aviation, cruise ship, etc.), and vector surveillance.

**Inject 3:**

One month later, it's September 2026, and several tourists are seeking medical care in Tropico, reporting fever, severe headaches, dizziness, and in a few cases, body rashes. Testing is limited, and samples are sent to a regional public health lab so that results take 10-14 days . Furthermore, following the International Football Championship, additional cases of riofiebre are identified in several countries in North America, Europe, Africa, and the Indo-Pacific. Due to the outbreak in San Pedro and Val Verde, many of these countries are beginning to integrate riofiebre into the traveler-based WES systems at airports.

So far, there are 23 confirmed and 8 inconclusive cases of riofiebre reported by the regional public health lab from the patient samples in Tropico. As a result of these cases, the recent attention to the outbreaks in San Pedro and Val Verde, travel-related cases in Pacifica, and many cruise-ships that frequent Tropico, health authorities have raised concerns about the limited surveillance capacity.

**Inject 3 Prompts:**

1. How should WES in isolated or rural areas, like Tropico, with considerable tourism/visitors be managed?
   - Would WES be a viable option?
   - What are the hurdles in establishing WES?
   - What are sustainable efforts that could be implemented?
2. Should the global community consider implementing WES at airports or other travel hubs like cruise ships to monitor wastewater from aircraft and airport facilities, offering a proactive or early detection system for riofiebre?
   - Consider the opportunity for flipping the WES model and conducting proactive surveillance rather than confirmatory surveillance.
   - Consider data integration from aviation with other sources of WES
3. What are the use cases for this aviation-focused data?
4. Could it be helpful in informing global surveillance initiatives and travel-related public health responses?
5. How could a proportionate response be considered?
   - What are the pros and cons from a Latin American perspective or sharing real time WES data with global surveillance initiatives?
   - Can the pros be maximized?
6. Given that the vectors transmitting riofiebre have established habitats in several of the countries and regions with post-International Football Championship cases, should these countries sustain surveillance targeting riofiebre?

***Supplementary Data 1: Panama GLOWACON Tabletop Event***

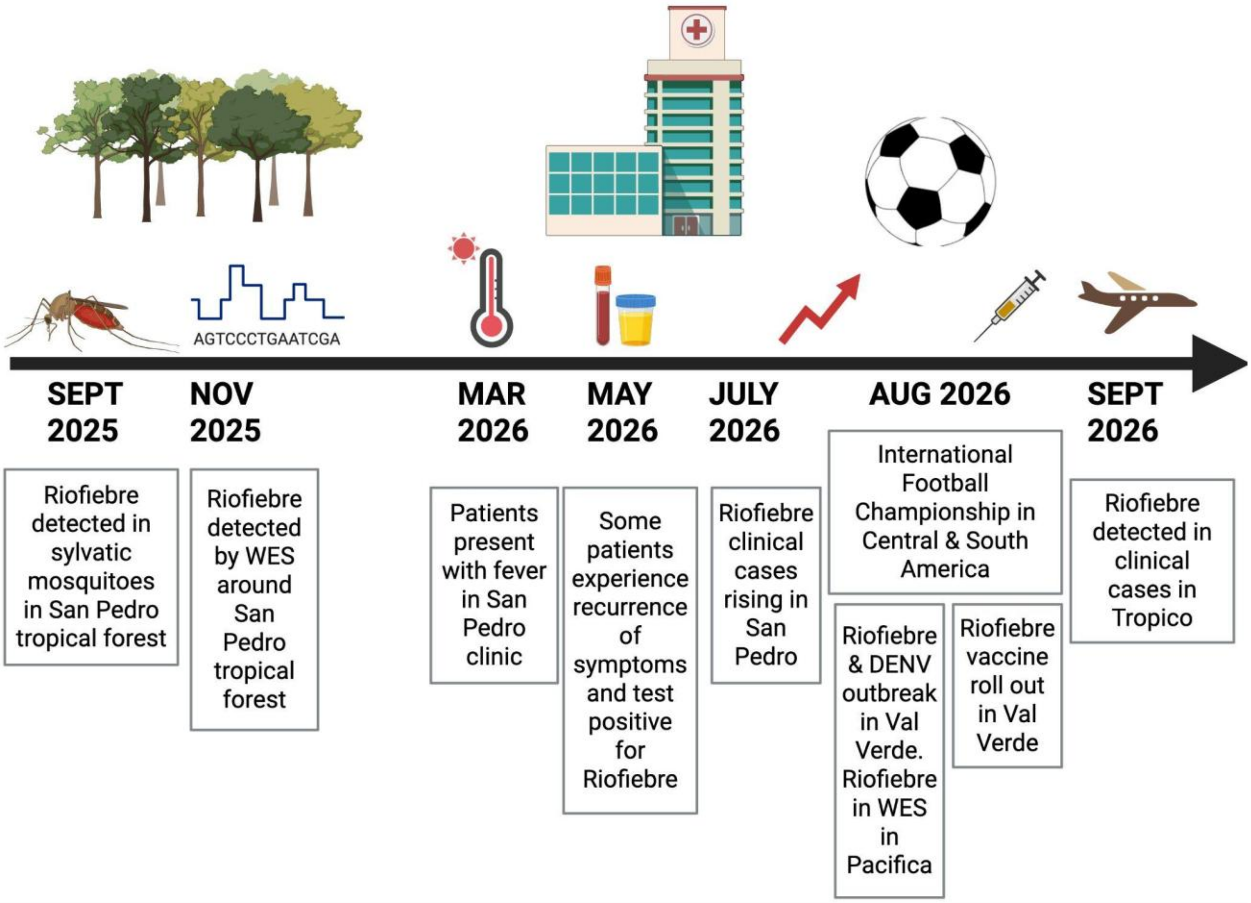

*Supplementary Data 1: Panama GLOWACON Tabletop Event*

### Post-event – Main Themes Identified

#### Base Scenario:

1. What was the significance of the wastewater and environmental monitoring finding and the level of concern it may warrant from a public health perspective?
   - Necessary to identify the source of the signal. *(3 groups, 4 responders)*
     - Animal matter, asymptomatic infected individuals, mosquito larvae
   - Important to establish an outbreak timeline. *(2 groups, 1 responder)*
   - Tools development to identify source. *(1 group, 1 responder)*
     - Sequencing, enteric markers, serological
   - Early warning useful to inform clinical partners to watch for symptoms. *(2 groups, 1 responder)*
   - Trigger vector surveillance; detection in vectors can indicate enzootic cycle. *(1 group, 1 responder)*
   - Low level of concern. *(1 group, 1 responder)*
   - Information sharing immediately important – PAHO, WHO, NGOs. *(2 groups)*

2. What should San Pedro public health officials do now that 40 more people in the tour group show laboratory evidence of riofever infection?
   - Declare an outbreak/epi alert. *(2 groups)*
   - Monitor infected persons and those close to them. *(2 groups)*
   - Monitor viral evolution and changes in fitness. *(2 groups)*
   - Targeted PCR testing. *(3 groups)*
   - Sequence pathogen. *(2 groups, 1 responder)*
   - Vector surveillance. *(2 groups, 3 responders)*
   - Increase WES efforts in neighboring areas. *(2 groups, 1 responder)*
   - Increase communication efforts with public and clinical community. *(2 groups)*
   - Increase lab and clinical capacities. *(2 groups)*
   - Conduct serosurveys to check if population has previously been infected. *(1 group, 1 responder)*

3. Explain the roles of the NGO conducting epidemiological surveillance in San Pedro, the San Pedro public health institute and PAHO/WHO at this point in the investigation.
   - NGO: genomics, testing, report findings to authorities. *(2 groups, 1 responder)*
   - Public health institute: analysis, detection, clinical investigation. *(2 groups, 1 responder)*
   - WHO: coordination, support. *(2 groups, 1 responder)*
   - PAHO: coordination, investigations, communication. *(2 groups)*

#### Inject 1:

1. What value, if any, would wastewater and environmental monitoring have in San Pedro, Val Verde, and Pacifica at this time?

- To generate baseline, early warning data. *(3 groups)*
- Use data for risk communication to inform citizens. *(3 groups)*
- Targeted WES at points of entry. *(1 group, 1 responder)*
- Test at event sites. *(1 group, 1 responder)*
- Increase vector monitoring. *(2 groups, 1 responder)*
- Need to validate WW detection ability based on sewage system. *(2 groups)*
- Depends on available system capacity. *(2 groups)*
- WES to determine disease incidence. *(1 group, 2 responders)*

2. What potential problems (and solutions) for implementing, sustaining and/or expanding wastewater and environmental monitoring in San Pedro and Val Verde should be considered?

- Financing would be a bottleneck. *(2 groups, 1 responder)*
    - Levy a tax on tourists to help fund
- Availability of testing reagents/lab equipment. *(2 groups, 1 responder)*
- Lab infrastructure/personnel. *(2 groups, 3 responders)*
- Targeted testing at points of entry. *(1 group, 2 responders)*
- Sample collection personnel capacity, training. *(1 group, 1 responder)*

3. How could sample collection, analysis and survey data best be integrated with existing surveillance systems and laboratory networks? What would collaboration with national and regional disease control centers look like?

- Integrate WES data with other surveillance data. *(3 groups)*
    - With existing dashboards, methods, communication networks
- Coordination with PAHO for regional support. *(2 groups)*
- Strengthen national/regional/international networks. *(2 groups)*
- Standardization and sharing of methods and data. *(1 group, 1 responder)*

4. Given what is known so far, what would be the communication plan for travelers, pregnant women, and physicians?

- Target communication at prenatal clinics, OBGYNs, pregnant women. *(3 groups)*
- Communicate clearly what is known and what is unknown. *(3 groups)*
    - Build confidence and aid individual decision making
- Communication across gov orgs to ensure a consistent and well-formed message. *(2 groups)*
    - Utilize a spokesperson
- Travel advisories and communication targeted at travelers. *(2 groups, 3 responders)*
    - Mosquito education, repellent, nets, appropriate clothing, etc.
- WES data not sufficient on its own, integrate with other data. *(2 groups, 1 responder)*

5. Would there be a value for doing aviation wastewater surveillance at this point- if so, which countries would benefit from this?
   - Especially useful for Pacifica and Val Verde. *(3 groups, 2 responders)*
     - Track returning travelers
   - Returning infected individuals would still reenter their countries, but still useful for tracking. *(1 group, 1 responder)*
   - Valuable for all involved countries. *(2 groups, 2 responders)*

6. What should the regional laboratory network consist of and what should be its priorities for establishing a response plan, taking into account the systems under stress and the burden of cases of riofever?
   - Should have lab testing capabilities. *(1 group, 1 responder)*
     - PCR, at least ability to refer for sequencing
   - Increase lab testing capacity/personnel. *(1 group, 1 responder)*
   - Regional collaboration/sharing of personnel and protocols. *(1 group, 3 responders)*

**Inject 2:**

1. What types of public health response actions could be triggered by the findings of wastewater and environmental surveillance both nationally and globally?
   - Increase population testing and vaccination events. *(1 group, 1 responder)*
   - Use WES data to enhance molecular epidemiology efforts to understand viral evolution, variants, pathogenesis. *(2 groups, 2 responders)*
   - Communicate findings at multiple levels and internationally. *(3 groups)*
   - Create agreements to collaborate and transfer resources between neighboring countries. *(1 group, 1 responder)*
   - Utilize lessons learned and communication channels developed during SC2 pandemic. *(2 groups)*

2. What additional wastewater and environmental surveillance research should be encouraged to optimize wastewater and environmental surveillance methods for detection of dengue and dengue fever and to better understand their molecular biology and epidemiology?
   - Virus stability and degradation rates. *(3 groups, 1 responder)*
   - Improve and standardize sampling and enrichment methods to increase sensitivity. *(2 groups)*
   - How to use WES data to evaluate and design more effective vaccines, diagnostics, treatment. *(3 groups, 1 responder)*
   - Analysis to understand viral evolution and variants. *(1 group, 1 responder)*

3. Should smaller, regional, non-case countries with minimal wastewater and environmental monitoring systems establish testing for riofever? Why or why not? What stronger recommendations would prompt them to implement wastewater and environmental monitoring?
   - It depends on country capacity and resource availability. *(2 groups)*
   - It depends if the pathogen/vector is already present. *(1 group, 1 responder)*
   - Scale up and down as needed based on regional alerts. *(1 group, 1 responder)*
   - Targeted, efficient WES sampling encouraged. *(2 groups)*
     - Airports, ports
     - Most populated areas
     - Academic institutions

4. Should there be additional actions around the soccer championship?
   - Yes, there definitely should be additional actions taken. *(3 groups)*
   - Issue warnings and guidance to participants. *(3 groups)*
   - Conduct vector control. *(2 groups)*
   - Enhance on-site surveillance and testing. *(3 groups, 3 responders)*
   - Don’t cancel event. *(2 groups)*

**Inject 3:**

1. How should wastewater and environmental monitoring be managed in isolated or rural areas, such as Trópico, with considerable tourism/number of visitors?
   - Limited sewer system makes WES challenging. *(3 groups)*
   - Monitoring tourist hubs using WES could be effective. *(2 groups, 2 responders)*
     - Cruise ships, hotels
   - Implement testing at key sites. *(3 groups)*
     - Airports, ports, transport corridors, bus stations
     - Sewage collection trucks in non-sewered areas
   - At this point, too late for WES – need to establish before outbreak. *(2 groups)*
   - Strengthen capacity, training, and support networks. *(1 group, 1 responder)*

2. Should the international community consider implementing wastewater and environmental surveillance at airports or other transportation hubs, such as cruise ships, to monitor wastewater from aircraft and airport facilities, providing a proactive or early detection system for river fever?

- Yes, the international community should implement WES proactively at transport hubs. *(4 groups, 3 responders)*
  - Cruise ships, airports, ports
  - Globally monitor returning population

3. What are the use cases for these aviation-focused data?

- Responsible data sharing to strengthen coordination but avoid panic. *(3 groups)*
  - Don't need to share raw, complete WES data
- Sharing of data showing disease spread negatively affects tourism. *(2 groups)*
- Can help improve preparedness and enhance national image. *(1 group, 2 responders)*
- Global sharing to aid tracking and response. *(2 groups, 1 responder)*

4. Given that the vectors that transmit riofever have established habitats in several of the countries and regions with cases following the International Football Championship, should these countries maintain targeted surveillance for riofever?
   - Yes, surveillance should be maintained. *(2 groups, 4 responders)*
   - Maintaining surveillance may be too costly for some countries. *(2 groups, 1 responder)*
   - It depends on disease seasonality and variant presence. *(1 group, 1 responder)*
