## Supplementary material for "How public health decision-makers operationalise wastewater surveillance: a multi-region qualitative study": Ethiopia Event S2

*Supplementary Data 2: Ethiopia GLOWACON Tabletop Event*

### Event Scenario & Prompts

Today's exercise takes place in fictitious countries in Africa and Europe. The disease is fictitious. This scenario takes place in 2025.

**Africa context:**

Country 1 is Wakanda, a country with highly sewered systems. WES in several areas being conducted by the government and NGOs.

Country 2 is Afrinia, a country with mixed sewage systems, some large cities have partial convergent sewered systems. Some WES being done by NGOs.

Country 3 is Ghudaza, a country with minimal convergent sewered systems. Some WES is being conducted for polio with support from NGOs.

Country 4 is Nadua, a country with minimal convergent sewered system. No WES underway yet.

**Europe context:**

Country 5 is Carpania, which has sewered systems and ongoing government run WES in community and airports.

**Base scenario:**

In March 2025, in Carpania in Europe, a new Mpox-like virus named X-pox is detected from clinical samples from three individuals who traveled from Nadua where they spent 4 weeks visiting family. The patients are two children ages 3 years and 5 years and their 26-year-old mother. X-pox illness is characterized by a high fever, headache, arthralgias, and a pox like rash that occurs on the face, mouth/throat, hands, trunk, and genitalia. All lesions appear to be at the same stage of development. The 26- year-old mother has a mild case with low grade fever and a few lesions; she is not hospitalised. The two children are admitted to the hospital and kept in isolation needing intravenous fluids, sedation, and pain relief. The 3-year-old develops uncontrolled seizures transferred to the ICU, intubated and remains comatose for 5 days and dies. IHR notifications are sent. The same day as the IHR notifications, officials in Wakanda report that an externally funded NGO doing WES pilot (that has not reported any pox virus to date) identifies X-pox in wastewater from multiple sampling locations. These WES X-pox detections are confirmed by a regional reference laboratory.

*Supplementary Data 2: Ethiopia GLOWACON Tabletop Event*

**Base Scenario Prompts:**

1. What is the significance of this WES finding?
   - What are the sources of the signal
   - How worrisome is this?
   - Does it warrant a response? By whom?
2. What next steps should Nadua (the country of origin of the case-patients) take?
   - What about data utilisation? What else do you need to know? What can you do while waiting?
   - What about further surveillance?
   - What about communication with key stakeholders and the public?
   - What are appropriate public health measures at this point that might be considered?
3. What should Wakanda (the country that detected X-pox in wastewater but does not yet have confirmed cases) consider doing next?
   - What about partnerships that may be required for expanded testing?
   - What challenges may you encounter in the absence of a clinical case?
4. What should the NGO doing WES pilot in Wakanda and Wakanda public health institute do together? What should the role of WHO and Africa CDC be?
   - What about data sharing and triangulation?
   - Consider communication
   - Consider ways of working
5. What collaborative or integrative surveillance measures should be implemented at this point? (please explore sample type, geographic region, etc.)

**Inject 1:**

Clinical cases of X-pox in children are reported from Afrinia and Ghudaza. These cases are more aggressive in children and in immunocompromised individuals, and show household transmission. In the capital city of Wakanda an outbreak of X-pox occurs in sex workers who seem to have milder disease. Wakanda public health sets up a testing program, but many adults in Wakanda refuse to be tested because of perceived stigma of the disease. Molecular biologists and phylogenetic experts have further characterized samples of X-pox and believe there are two variants corresponding to the milder form in adult sex workers and the aggressive severe form in children and immunocompromised adults.

**Inject 1 Prompts:**

1. What value, if any, would WES have in Afrinia and Ghudaza at this point?
   - What are the factors that might dictate this?
2. What are some potential problems and solutions for implementing WES in Wakanda, Nadua, Ghudaza and Afrinia?
   - Are there any capacity bottlenecks?
   - Are there any infrastructure issues?

- What can be done to resolve issues, if any?

3. How would you quickly and effectively build capacity to empower local personnel with WES methodologies, sample collection, processing, and data interpretation?

**Inject 2:**

Six months later, the outbreaks are ongoing in Wakanda, Nadua, Ghudaza, and Afrinia, and case reports are increasing. Two world renowned institutes in Africa and Europe report on a novel wastewater sampling processing method that enables high efficiency, high depth sequencing of X-pox and the two variants in wastewater. Simultaneously a global development organisation sets aside a large amount of money to set up WES surveillance across Africa. A vaccine company develops, tests a new vaccine at lightning speed, but supplies are limited. Another global charity is set to distribute vaccines where needed most.

**Inject 2 Prompts:**

1. How would we best integrate WES data with existing surveillance systems?
    - Collaboration with national and regional disease control centres
2. What types of public health response actions could be triggered by WES findings both nationally and globally?
    - Consider investigation and enhanced surveillance
    - Consider medical and non-medical interventions
    - Consider engagements across government and research stakeholders
    - Consider public communication
    - Consider international stakeholder engagement
3. What additional WES research should be encouraged to optimize WBE methods for X-pox detection and to better understand its epidemiology?
    - Consider the virus's persistence and viability in wastewater
    - Consider trials?

**Inject 3:**

A few more months go by, and additional severe paediatric cases are detected in travellers to multiple global destinations including in North America, Asia and the Middle East. Further expansion of cases are also detected across land borders in Africa.

**Inject 3 Prompts:**

1. Should the global community consider implementing WES at airports to monitor wastewater from aircraft and airport facilities, offering a proactive or early detection system for X-pox?
  - Consider the opportunity for flipping the WES model and conducting proactive surveillance rather than confirmatory surveillance.

- Consider data integration from aviation with other sources of WES

2. What are the use cases for this aviation-focused data?
    - Could it be helpful in informing global surveillance initiatives and travel-related public health responses?
    - How could a proportionate response be considered?
    - What are the pros and cons from an African perspective or sharing real time WES data with global surveillance initiatives?
    - Can the pros be maximised?

### Post-event – Main Themes Identified

#### Base Scenario:

1. What is the significance of this WES finding? What are the sources of the signal? How worrisome is this? Does it warrant a response? By whom?

Overall, the groups found the WES finding significant and worrisome, particularly due to the IHR notification, the geographic span, and the fatality. The most common warranted response was in country surveillance with international support/funding.

- 6 groups noted that this finding should specifically trigger increased clinical testing or syndromic surveillance/epidemiological mapping
- 3 groups specified that this should trigger more environmental testing.
- 3 groups specified that support should come from international networks (such as WHO).
- 3 groups noted that the finding could indicate either asymptomatic spread (1 group) or an animal reservoir (2 groups).
- 2 groups recommended ports of entry or travellers should be screened from now on.
- 2 groups noted the fatality as particularly worrisome.

2. What next steps should Nadua (the country of origin of the case-patients) take?

What about data utilisation? What else do you need to know? What can you do while waiting? What about further surveillance? What about communication with key stakeholders and the public? What are appropriate public health measures at this point that might be considered?

Groups wanted to know more about the pathogen – sequence results, transmission mode, incubation mode, etc. Groups focused on contract tracing, supporting hospitals and clinics, and forming effective government response teams to implement public health measures. There was also a strong push to strategically expand WES at this time.

- 4 groups stressed public risk communication, with 2 other groups stressing academic/public health notifications
- 4 groups wanted to expand WES into hotspots or travel hubs (including airport WES)
- 4 groups specified they wanted to activate emergency protocols in some way – rapid response teams, emergency plans, Task Force, etc.

- 5 groups wanted to focus on contract tracing
- 4 groups wanted to provide education and support to affected hospitals and clinics
- 3 groups wanted a concrete case definition

3. What should Kimbala (the country that detected X-pox in wastewater but does not yet have confirmed cases) consider doing next?

What about partnerships that may be required for expanded testing? What challenges may you encounter in the absence of a clinical case?

There was not a lot of overlap on specific actions or challenges facing Kimbala, but the overall theme was that Kimbala should widen the net and search for cases, increase data sharing/knowledge of the pathogen, and prepare public health response agencies.

- 3 groups wanted to expand environmental surveillance, specifically at airports and other ports of entries
- 3 groups wanted to expand awareness of a potential outbreak, either by looping in governmental agencies, private sectors industries, or general population education.
- 2 groups wanted to start clinical surveillance
- 2 groups wanted to expand data exchange with other international partners (WHO and Africa CDC cited specifically)
- Challenges included a lack of trust in health authorities (2), contact tracing if WES is deployed in a large sewer shed, and limited testing capacity.

4. What should the NGO doing WES pilot in Kimbala and Kimbala public health institute do together? What should the role of WHO and Africa CDC be?

What about data sharing and triangulation? Consider communication. Consider ways of working.

NGO:

- 5 groups thought that the NGO needed to share their information (both nationally and internationally)
- 3 groups though they should aid in increasing testing (lab) capacity and resource procurement
- 2 groups thought they should expand WES

Africa CDC/WHO

- 2 groups though their main role was to create an information sharing platform
- Other potential responsibilities included general support, protocol creation, and communication/media interaction.

5. What collaborative or integrative surveillance measures should be implemented at this point?

Please explore sample type, geographic region, etc.

2 groups did not respond to this question

- 5 groups (all that responded) wanted to expand surveillance.
- 2 groups wanted to focus on clinical surveillance
- 2 groups wanted to have a cross-border focus (including airport wastewater surveillance (2) and information sharing among countries)
- 2 groups wanted to implement contact tracing.
- One group wanted a coordinated One Health approach (including livestock and increased environmental surveillance)

**Inject 1:**

1. What value, if any, would WES have in Afrinia and Ghudaza at this point?

What are the factors that might dictate this?

- 5 groups thought WES would provide significant value through genomic sequencing and identification of new strains
- 3 groups thought WES would provide better epidemiological data, including prevalence (3), geographic location (allowing for targeted hotspot surveillance) (3), and trend monitoring over time (2).
- 2 groups thought that WES would catch cases that would otherwise go unreported due to testing stigma.
- Other values cited included filling the gaps of clinical surveillance, early detection of mild/asymptomatic cases, and the low cost.
- Factors that dictate WES's value included: population coverage and testing capacity.

2. What are some potential problems and solutions for implementing WES in Kimbala, Nadua, Ghudaza and Afrinia?

Are there any capacity bottlenecks? Are there any infrastructure issues? What can be done to resolve issues, if any?

Again, there was not a lot of overlap of the answers here (except for the worry over stigma).

- 4 groups expressed concern over the risk of stigma and/or community resistance to WES implementation.
- Capacity bottlenecks included: sampling capacity, resource procurement concerns, cost of transportation.
- Infrastructure issues include: lack of lab space, lack of effective sewage systems (Nadua specifically), and decentralized wastewater systems (listed as different than lack of).

- Additional problems cited were concern over the ability to interpret the collected data into effective public health actions and how to prioritize limited resources.
- Solutions were: secure more funding, scale up WES, build up capacity, leverage existing networks, and alternate sampling locations in decentralized systems.

3. How would you quickly and effectively build capacity to empower local personnel with WES methodologies, sample collection, processing, and data interpretation?

Groups focused on expanded knowledge through training programs, implementing international partnerships, and effective communication of WES data.

- 5 groups specified they would either follow international guidance or request direct support from WHO, Africa CDC, or Glowacon.
- 5 groups focused on training local staff and implementing a cascading chain of training (peer to peer, etc.)
- 3 groups mentioned increasing the effectiveness of data interpretation through training, visualization, and incorporation with clinical data.
- 2 groups would rely on targeting WES sampling of hotspots or large gathering areas.
- 2 groups would lobby for both money and capitol to build capacity, raising pollical awareness as well.
- Other noteworthy responses included preparing pathogen agnostic training plans, building on polio progress, and ensuing an effective supply chain.

**Inject 2:**

1. How would we best integrate WES data with existing surveillance systems?

Collaboration with national and regional disease control centres

- All seven groups wanted to integrate WES with clinical case data, often citing the creation of a visually appealing dashboard to do so.
- 3 groups wanted to integrate with vaccine data to inform vaccination policy.
- 2 groups wanted to put out weekly or monthly reports of the aggregated data.
- 2 groups wanted to use WES to identify hot spots where more clinical testing should be performed.
- 2 groups wanted to share WES data with regional, national, and other partners (“scientific advisory board”).

2. What types of public health response actions could be triggered by WES findings both nationally and globally?

Consider investigation and enhanced surveillance. Consider medical and non-medical interventions. Consider engagements across government and research stakeholders. Consider public communication. Consider international stakeholder engagement.

- 2 groups did not respond
- 5 groups thought the WES data could be used to guide vaccination policy as well as monitor the efficacy of vaccination strategies (ring vaccination, etc.)
- 5 groups wanted to use the WES findings to trigger risk communication to the public, raising civilian awareness.
- 2 groups though WES could trigger resource prioritization to hotspots (both in terms of location and demographics).
- 2 groups thought WES could trigger behavioural change PH actions (social distancing, increased hygiene, etc.)
- 2 groups thought WES could trigger travel restrictions (and guide their implementation) and trigger increased airport surveillance.
- Other noteworthy actions including expanding WES, triggering lab confirmation of clinical cases, and increased education of healthcare personnel.

3. What additional WES research should be encouraged to optimize WBE methods for X-pox detection and to better understand its epidemiology?

Consider the virus's persistence and viability in wastewater. Consider trials?

- 1 group did not respond
- 5 groups wanted to conduct trials (formally or informally) on how to optimize WES (by studying sampling method, viral degradation kinetics, transmission modes, etc.)
- 4 groups wanted to research shedding rates and viral persistence.
- 3 groups wanted to research vaccine efficiency (some tying this to shedding rates).
- 3 groups wanted to research variants and NGS data.
- Other research areas included linking WES to clinical correlates, mapping sewer systems with population density, and performing a cost benefit analysis. One groups stressed the need to create an appealing dashboard and visuals to convey the data.

***Supplementary Data 2: Ethiopia GLOWACON Tabletop Event***

**Inject 3:**

1. Should the global community consider implementing WES at airports to monitor wastewater from aircraft and airport facilities, offering a proactive or early detection system for X-pox?

Consider the opportunity for flipping the WES model and conducting proactive surveillance rather than confirmatory surveillance. Consider data integration from aviation with other sources of WES.

- 3 groups answered YES, 4 groups were unclear or conditional
- 3 groups cited the risk of unintended economic or reputational risk.
- 2 groups did not think WES should be freely at airports – it should either not be tied to the actual aircraft and rather be community wide, or it should be very targeted on certain aircraft.
- While 2 groups reiterated that WES was proactive, another group mentioned that it is still generally seen as reactive as it is a public health measure. This in turn increases the risk of reputational harm and/or public health panic.
- Other interesting comments here were that it could be used only for variant monitoring, it will present challenging data interpretation, it is cost efficient, and would provide a wealth of information. Overall, very mixed answers, but no outright "NO"s.

2. What are the use cases for this aviation-focused data?

Could it be helpful in informing global surveillance initiatives and travel-related public health responses? How could a proportionate response be considered? What are the pros and cons from an African perspective or sharing real time WES data with global surveillance initiatives? Can the pros be maximised?

- 4 groups did not respond
- The 3 groups that did respond all thought it would be helpful to global surveillance and provide helpful data.
- 2 groups stressed the importance of anonymized data, one group stressing that it should be deployed at the level of the airport, not the aircraft.
- 2 groups stressed how important WES could be for early detection. Other Pros included variant detection and another piece of data to make sense of other data (clinical surveillance, symptom reports, etc.)
- Cons included reputational risk, a lack of public understanding on WES data implication, and the idea of missing data from short flights.
- Pros could be maximized by deploying WES during large international events (i.e., Olympics).
