## Supplementary material for "How public health decision-makers operationalise wastewater surveillance: a multi-region qualitative study": Singapore Event S3

*Supplementary Data 3: Singapore GLOWACON Tabletop Event*

### Event Scenario & Prompts

**Base Scenario:**

On April 15, 2025, a veterinary hospital in North America reported caring for 5 cats who succumbed to a respiratory infection. All routine diagnostic tests were negative and the national animal agriculture agency of that country completed additional tests and issued an alert that a highly pathogenic avian influenza (HPAI) virus was the cause of the illness. The cat owners were interviewed, observed, and tested but additional information was not available.

You are on a multidisciplinary team put together by your government. Some decisions need to be made immediately regarding this potentially new HPAI that has emerged with no obvious common source. You are responsible for advising on the use of WES.

**Base Scenario Prompts:**

1. Why would or wouldn't you use WES at this time?
2. What information would you want to know to drive your decision to start WES?
3. What gaps in surveillance would you fill through use of WES?
4. What are the pitfalls and limitations of using WES at this time?

**Inject 1:**

On June 25, 2025, 6 days into a 10-day bird watching trip in North America, a 65-year-old husband and his 68-year-old wife became ill with a cough and fever of 39 C that was getting worse, so they went to see the clinician at a local urgent care. During the visit the husband began complaining of chest pain and shortness of breath and was admitted to the hospital via ambulance and required supplemental oxygen. The couple were up to date on their adult immunizations and had received flu and COVID boosters in October 2023. They were avid travelers who lived in a suburban area with two cats, one of whom died immediately prior to their trip. They travel globally 3-4 times per year for bird watching and other nature trips. On this trip they stayed at a lodge located on a cattle ranch where there were 15 other guests and two guides. The lodge had 15 employees who were temporary workers who arrived at the ranch from Central America in September. Nasal swabs samples for influenza and SARS-CoV-2 were taken.

**Inject 1 Prompts:**

1. Why would or wouldn't you use WES at this time?
2. What information would drive your decision to start WES?
3. What gaps in surveillance would you fill through use of WES?

4. What are the pitfalls and limitations of using WES at this time?

**Inject 2:**

On June 30, 2025, the Ministry of Health in a country in Asia reports, via an IHR notification, that a 19-year-old University student who recently returned home from a Semester at Sea program was diagnosed with a respiratory illness and testing revealed a highly pathogenic avian influenza strain not previously found in humans. During the semester at sea the student complained of a respiratory illness like a “cold” that many of his classmates had too. The symptoms included headache and a sore throat. On exam he had a low-grade fever. Throat cultures for Group A Strep and SARS-CoV-2 testing were negative. The summer semester at sea began in May 2024 and lasted 106 days until late August 2024. It included stops in 10 countries and 11 cities on 3 continents (Africa, Asia and Europe).

On July 4, 2025, a 40-year-old international airport worker in a major metropolitan city in North America presented to healthcare with a fever of unknown origin and mild respiratory symptoms for 2 weeks. Local public health authorities were notified. She did not travel but works in a spa concession at the airport. She lives at home with her elderly mother and is a fitness buff who often drinks unpasteurized (raw) milk that she buys from a dairy farm in a rural area two hours from the city. She is <u>not</u> up to date on her adult immunizations and cannot recall receiving an influenza vaccine for at least 20 years; she has not had any COVID-19 vaccines. Testing reveals that she has HPAI (same strain as reported in the university student).

On July 5, 2025, the lab results for the couple on the birdwatching trip indicate that they have HPAI (same strain identified in the student and the airport worker).

**Inject 2 Prompts:**

1. Why would or wouldn’t you use WES at this time?
2. What information would drive your decision to start WES?
3. What gaps in surveillance would you fill through use of WES?
4. What are the pitfalls and limitations of using WES at this time?
5. Which populations would you target? Would you use airplane/airport WES? Why?
6. Who would be relevant partners in all levels of government and in other sectors (i.e., industry, research/scientific community, third-party) and what are the outcomes that each partner group might be concerned about?
7. Who would you share the data with? And for what purpose? (e.g. will data integration occur with other surveillance data to triangulate with other data and analysis or who will need the data for decision making /response)
8. What other information might you need to consider in order to utilise WES or derive added value from it?

*Supplementary Data 3: Singapore GLOWACON Tabletop Event*

**Inject 3:**

It is now three years later, and the disease is declared endemic with no clear seasonal pattern.

**Inject 3 Prompts:**

1. How will you adjust your WES strategy to address this issue?
2. What changes would be made?

**Inject 4:**

You have been asked to advise the public health ministry in a lower income country with limited resources. You are asked to assess the use of WES in this LMIC for the same HPAI pathogen that is now endemic.

**Inject 4 Prompts:**

1. What are your recommendations?
2. Why would or wouldn't you use WES at this time?
3. What information would drive your decision to start WES?

### Post-event – Main Themes Identified

**Base Scenario:**

1. Why would or wouldn't you use WES at this time?
    - 10 of the response groups would use WES, 5 would not, and 4 were either unclear in their response or indicated that they were undecided.
    - "No" responses centered on the lack of available epidemiological information, including transmission dynamics and the virus's infectivity in humans. "No" answers were uncomfortable justifying the expense without a better indication that it would be worthwhile.
    - "Yes" responses were often contingent on having existing capacity. Many wanted to deploy WES in a limited capacity to begin with, including to pinpoint the source of the outbreak.
    - Both YES and NO responses often referenced how little data they had to work with. To resolve the lack of information, answers were split between 1) *using* WES to gain that information and 2) waiting for it to come from other sources.

2. What information would you want to know to drive your decision to start WES?

The most commonly requested information was:

- 10 groups wanted to know if there was circumstantial evidence of human infection (similar symptoms, odd flu-like illnesses, etc.). These answers were searching for evidence of human transmission beyond a concrete confirmatory laboratory test.

- 8 groups wanted explicit epidemiological data of feline cases, including links between cases, the timeline of infection, potential avian exposure, and environmental data from wild animals.
- 5 groups wanted to know the current resources of the area, and if WES capacity already existed.
- 5 groups wanted to know the genomic sequence, with one answer citing that the genome could be used to predict the risk for potential zoonotic transmission.
- 4 groups wanted expanded test results, on both potential human exposures and the sickened cats. Confirmatory diagnostic tests were often cited as a *requirement* to move forward with WES implementation, of both the cats and the owners.
- 3 groups wanted explicit epidemiologic data of human cases if they were identified, including geospatial distribution of cases, transmission patterns, and health and travel history of cases.

3. What gaps in surveillance would you fill through use of WES?
    - 13 groups wanted to establish the distribution of the disease and determine how widespread the infection currently is. Respondents wanted to use WES to build an epidemiologic map and identify where the infection was circulating.
    - 5 groups wanted to determine if asymptomatic transmission was occurring.
    - Some respondents wanted to use WES to identify the source of infection, although other groups did not think it could accurately determine the source.
    - 3 groups cited the use of WES to supplement missing syndromic surveillance and to use it to validate clinical data.

4. What are the pitfalls and limitations of using WES at this time?
    - 6 groups responded that at this time, the test would have to be quite sensitive in order to pick up the likely low level of virus in the water, and even then, it may not be sensitive enough. Some groups mentioned that a negative result would not certify an area HPAI-free, similarly, a positive result may cause undue alarm.
    - 5 groups were uncomfortable using WES when there was not enough information specifically on the best geographic site for sampling.
    - 4 groups were uncomfortable using WES due to a knowledge gap in general, meaning that the lack of contextual epidemiological data meant the results may not be interpreted correctly.
    - 4 groups thought that the inability to distinguish between animal and human cases was a significant pitfall
    - 4 groups thought that the cost and potential waste of resources, particularly if WES capacity was not already present, was a significant pitfall.

**Inject 1:**

1. Why would or wouldn't you use WES at this time?

- 3 groups would use WES, 13 groups would NOT, and 3 were undecided. Not all groups recorded their reasoning
- Of the groups who would not use WES, 9 groups wanted to gather more epidemiological data before initiating WES. Groups wanted to collect data from traditional sources first, including testing all exposed individuals and initiating contact tracing/case finding. Contact tracing and clinical surveillance were strongly preferred over initiation of WES.
- 2 groups would not use WES because the causative agent was not yet identified.
- 2 groups who indicated they were undecided would use WES if a case was found during traditional epidemiological investigations or if resources (money) was left over after individual testing was completed.
- Of the groups who would use WES, 1 would do so in a limited, focused capacity, while the other group that explained their reasoning thought WES was the most useful when it was continuously implemented.

2. What information would you want to know to drive your decision to start WES?
   - By far, the most common response was to know the results of the tests. 15 groups requested the results of the nasal swabs.
   - 9 groups wanted further epidemiologic and clinical data, including the couple's travel history, symptoms of other lodge guests, geographic distribution of any symptomatic individuals, and whether there has been an uptick in flu-like illness in the local community. Groups wanted to know whether there was contact tracing and/or active case finding currently ongoing.
   - 3 groups wanted further information on the cat and a definitive cause of death
   - 3 groups wanted genomic data from the test results.
   - Further requested information (each had 2 groups mentioning it) were: whether there was a linkage between the cats in the previous section and the couple, results of wastewater collection, and the status of resources available to public health efforts.

3. What gaps in surveillance would you fill through use of WES?
   - 8 groups wanted to use WES to determine how widespread the virus may be in the region.
   - 2 groups wanted to use it to locate the source of infection.
   - 2 groups wanted to use the data generated to build predictive epidemiologic models.
   - 2 groups would no fill in any gap – because they were adamant about not using WES.
   - Other interesting gaps cited were: to look for asymptomatic cases and to investigate the existence of a reservoir in wild animals.

4. What are the pitfalls and limitations of using WES at this time?

- The most common limitation a concern that the lack of direct evidence of an infection would create an inefficient use of resources at this stage in the investigation. 11 groups expressed this concern in one form or another, citing issues with the lack of information (unknown source of couple's infection, the couple's and other lodge guest's transitory nature, the cost of community wide WES implementation, etc.). 2 groups were concerned specifically with the lack of information in relation to when and where WES sampling would even occur.
- 3 groups expressed concerns over the sensitivity of WES.
- 3 groups expressed concern over the inability to distinguish between animal and human cases

**Inject 2:**

1. Why would or wouldn't you use WES at this time?
   - Finally – every group (19) would use WES!
   - 5 groups would use WES now that there is not an information gap: the causative agent has been identified and overall there is enough information from traditional sources that they feel confident utilizing WES is not a overreaction.
   - 4 groups would use WES now that the virus is clearly widespread and is no longer confined to obvious epidemiologic linkages.
   - 2 groups would only use WES provided there is existing capacity.

2. What information would you want to know to drive your decision to start WES?
   - Answers were significantly more spread out for this question than previous questions.
   - 5 groups wanted to know the geographic location of cases in order to prioritize sampling and/or to investigate epidemiological links.
   - 4 groups wanted to know the genomic sequence.
   - 3 groups wanted to ensure that they had a pre-tried assay that they knew would work, and that capacity existed in country prior to WES implementation.
   - 3 groups wanted more clinical surveillance data, and wanted to know how to relate this type of traditional data to WES.
   - 3 groups wanted evidence of human-to-human transmission, with one group wanting evidence of community transmission.
   - The following information had 2 groups each indicate they would want to know:
     - That the cases indeed were all the same HPAI
     - The number of international cases
     - The severity and symptoms of the illness.

3. What gaps in surveillance would you fill through use of WES?

- The most common response (13 groups) by far was to use WES to gather data on the prevalence of the virus in the population and the geographic distribution of cases. Groups were looking for evidence of community transmission
- 4 groups would use WES to learn more about asymptomatic cases.
- 3 groups would use WES to learn how the virus's genome evolves over time.
- 3 groups would use WES as a supplement to clinical surveillance.
- 3 groups would use WES as a monitoring tool to be used in real time to adjust public health measures as needed.

4. What are the pitfalls and limitations of using WES at this time?
    - 5 groups were concerned with the information that WES can NOT provide, including disease severity, whether the virus was shed from an animal or a human, and the demographics of the human cases.
    - 4 groups were concerned with the cost of WES implementation and a potential waste of resources, including the time needed to create an assay.
    - 4 groups were concerned with the global issues surrounding WES implementation, and that there were too many countries trying to coordinate.
    - 3 groups were concerned that the shedding rate of the virus was not known, and how that would impact WES's accuracy.
    - 2 groups were concerned about potential urban bias due to a lack of rural infrastructure.

5. Which populations would you target? Would you use airplane/airport WES? Why?
    - Several groups only answered 1 part of this 3-part question (and it varied which part they chose to respond to). Of the responses received:
        - 1 group would not use airport WES, 1 group was split amongst themselves, and 11 groups would use it.
        - Targeted populations included anyone using the airport (groups often did not specify workers vs travelers but just wrote airport – 7 groups), residents of urban areas (7), international travelers (distinguished from the 'airport' answer – 4 groups) the cruise ship and/or the locations visited (3), contacts identified from contact tracing (3), university students (2), and local, high-risk populations (2).
    - Those who would use airport WES and gave a reason for doing so often cited the richness of the data compared to sewage WES. It is much more specific and can be used to easily trace where the infected individual arrived from. This helps establish geographical links.

6. Who would be relevant partners in all levels of government and in other sectors (i.e., industry, research/scientific community, third-party) and what are the outcomes that each partner group might concerned about?

- Groups did not always specific the outcome they expected from these partnerships, but the most common answers that were written are included below.
- 6 groups would want to partner with the Environmental Department at various levels of government, as they are likely the branch of government that handles wastewater.
- 6 groups would want to partner with public health departments at the federal level of government/Ministries of Health.
- 6 groups cited using a 'whole of government approach' (specifically written in one response), intended to balance competing interests and capture more viewpoints.
- 6 groups would partner with the scientific community and/or national reference labs. These partners would be concerned about learning as much as possible about the virus, including transmission pathways, high-risk populations, and potential vaccination strategies.
- 6 groups would partner with industry leaders in general (as opposed to the aviation industry specifically) to secure resources and the necessary supply chains. This group was cited as being concerned with economic losses.
- 5 groups would want to partner with public health professionals doing clinical surveillance work, as they are concerned with generating epidemiological data and stopping the outbreak.
- 4 groups would partner with either the WHO or CDC/country CDC equivalent.
- 3 groups would partner with the aviation industry, as they would be concerned about mitigating potential economic loss.
- 3 groups would partner with the agriculture industry, as this group is concerned with the protection of stock (good for One Health applications at disease prevention) and economic loss from a zoonotic disease.

7. Who would you share the data with? And for what purpose?
   - The overarching response was that data should be shared, but the raw data should only be shared in-country and potentially with the WHO – all other sharing should of analysis and interpretation only.
   - 11 groups specifically mentioned sharing with governments, from the local level up to Ministries of Health, for outbreak response.
   - 6 groups wanted to specifically share with the WHO and other international organizations, again for outbreak response.
   - 4 groups mentioned sharing with academia and the larger scientific community, to understand the disease and to develop therapeutics.
   - 2 groups advocated for a public dashboard to be made available to all, to easily track trends in cases.

8. What other information might you need to consider in order to utilize WES or derive added value from it?

- 8 groups wanted to know the clinical course of the disease, including how severe the illness is in humans. One group specifically wanted to know the outcome of feline cases as well.
- 6 groups wanted detailed epidemiological data derived from contact tracing and active case finding.
- 4 groups wanted to know the global status of the outbreak, for added context when determining if they should continue to use WES.
- 3 groups wanted the genomic sequence of the virus.
- 3 groups wanted more information on viral shedding, including shedding rates.
- 3 groups wanted more detailed information of transmission dynamics and infectivity of the virus.
- 2 groups wanted to know whether a vaccine would soon become available.
- 2 groups wanted to know more about the wastewater and sewage system layout where WES was deployed.

**Inject 3:**

1. How will you adjust your WES strategy to address this issue?
    - 12 groups would continue to utilize WES, but at a less frequent rate (ranging from weekly to monthly or to when prompted by clinical surveillance data).
    - 7 groups wanted to scale back WES and to deploy it in focused geographical areas. Groups wanted to link the use of WES to hotspots discovered in clinical data.
    - 7 groups wanted to use WES to surveil for variants of the virus. One group wanted to know the severity of resulting variants in order to determine if they would continue to use WES.
    - 2 groups would stop using WES once the virus became endemic, with one of these groups doing so slowly as they adjusted based on historical data.
    - 2 groups wanted to take WES national at this stage.
    - 2 groups wanted to continue WES as long as the cost was low/justifiable.
    - 2 groups wanted to evaluate if WES could be expanded to other novel pathogens in order to justify its continued use.

2. What changes would be made?
    - 11 groups would adjust the frequency of the sampling. Most of these groups would keep use WES at a regular interval, but the frequency would be scaled down.
    - 7 groups would adjust the location of the sampling as well, focusing on hotspots identified through clinical surveillance and other tools.
    - 6 groups would keep track of genomic data to monitor for new variants of the virus.
    - 5 groups would focus on making WES sustainable by lowering the cost and/or using it for other pathogens to make continuous use more worthwhile
    - 2 groups would focus on developing and contributing to a global network to facilitate data sharing.

- 2 groups would pursue further research to understand transmission dynamics to better utilize WES.

**Inject 4:**

1. What are your recommendations?
    - 8 groups would use WES in a targeted, informed manner by lowering the frequency and choosing specific sampling sites rather than national surveillance. Answers suggested using historical data, limiting surveillance to areas with sewage systems, and using clinical data to guide sampling sites.
    - 6 groups would partner with countries that have experience using WES, with some suggesting the development of a regional network to support LMIC with WES implementation.
    - 3 groups would invest in R&D, to develop tools such as vaccines, therapeutics, and cheaper diagnostics.
    - 3 groups would stress that funding will be made available to help with implementation.
    - 2 groups would conduct pilot studies of WES in hotspots to prove its worth.
    - 2 groups would embark on a public information campaign to stress the benefits to locals.

2. Why would or wouldn't you use WES at this time?
    - 12 groups would use WES, 4 might depending on circumstances, and 2 would not. One group did not answer this question.
    - For those that would use WES, it often came with caveats, including:
        - If capacity was already robust
        - Use only on a limited, as-needed basis
        - Use at a reduced frequency
    - 6 groups would use WES to provide either additional contextual data or in place of surveillance data, in order to maintain a baseline and detect a change in caseload.
    - 5 groups would use WES because of the low cost, with tweaks to the protocol in order to make it even more cost effective (such as broad coverage surveillance).
    - 4 groups would use WES to ensure that the capacity for use remained intact. Some groups specified that they would help build capacity so the country was better prepared for future infectious disease outbreaks and was able to detect other pathogens.
    - 2 groups would have WES on a standby basis, activating if clinical surveillance data showed an increase in disease.
    - For those who might use WES (those groups that either did not provide a clear answer or gave reasons for both), capacity and cost were the two most common considerations. 3 groups would use WES if the cost could be justified by better health outcomes in the population.

- For those who would not use WES or might not, 2 groups would not do so because of the cost – they believed limited resources could be channeled to higher priority targets. Other answers included the redundancy if traditional clinical surveillance data was already robust and if the sewage infrastructure was not widespread and well-maintained.

3. What information would drive your decision to start WES?
    - 5 groups wanted information on the sewage system and the population distribution in relation to said system, in order to ensure WES would capture enough of the population.
    - 5 groups wanted to ensure the public health infrastructure was robust enough in-country to be able to both analyze and effectively use the data generated from WES.
    - 5 groups wanted to know the feasibility of WES use in-country, including community buy-in and technical capacity.
    - 4 groups wanted to use WES if clinical rates indicated a surge in cases, or if there was a gap in clinical data that WES could help fill.
    - 4 groups would need to know that funding was secured in order to start WES, with one group specifying that WES implementation must be cheaper and add more value than the current surveillance system in place.
    - 3 groups wanted to know the impact of the pathogen on the population, including its severity and transmissibility.
    - 2 groups wanted to know the regional trends in neighboring countries.
